# Natural Language Model for Automatic Identification of Intimate Partner Violence Reports from Twitter

**DOI:** 10.1101/2021.11.24.21266793

**Authors:** Mohammed Ali Al-Garadi, Sangmi Kim, Yuting Guo, Elise Warren, Yuan-Chi Yang, Sahithi Lakamana, Abeed Sarker

## Abstract

**Background:** Intimate partner violence (IPV) is a preventable public health issue that affects millions of people worldwide. Approximately one in four women are estimated to be or have been victims of severe violence at some point in their lives, irrespective of their age, ethnicity, and economic status. Victims often report IPV experiences on social media, and automatic detection of such reports via machine learning may enable the proactive and targeted distribution of support and/or interventions for those in need.

**Methods:** We collected posts from Twitter using a list of keywords related to IPV. We manually reviewed subsets of retrieved posts, and prepared annotation guidelines to categorize tweets into *IPV*-*report* or *non-IPV-report*. We manually annotated a random subset of the collected tweets according to the guidelines, and used them to train and evaluate multiple supervised classification models. For the best classification strategy, we examined the model errors, bias, and trustworthiness through manual and automated content analysis.

**Results:** We annotated a total of 6,348 tweets, with inter-annotator agreement (IAA) of 0.86 (Cohen’s kappa) among 1,834 double-annotated tweets. The dataset had substantial class imbalance, with only 668 (∼11%) tweets representing IPV-reports. The RoBERTa model achieved the best classification performance (accuracy: 95%; IPV-report F_1_-score 0.76; non-IPV-report F_1_-score 0.97). Content analysis of the tweets revealed that the RoBERTa model sometimes misclassified as it focused on IPV-irrelevant words or symbols during decision making. Classification outcome and word importance analyses showed that our developed model is not biased toward gender or ethnicity while making classification decisions.

**Conclusion:** Our study developed an effective NLP model to identify IPV-reporting tweets automatically and in real time. The developed model can be an essential component for providing proactive social media based intervention and support for victims. It may also be used for population-level surveillance and conducting large-scale cohort studies.

## INTRODUCTION

Intimate partner violence (IPV) is a preventable public health issue that seriously affects millions of people worldwide [1]. IPV can be defined as physical or sexual assault, or both, of a spouse, partner, or cohabiting dating couples [2, 3]. Approximately one in four women in the United States (US) are estimated to be or have been victims of severe violence at some point in their lives, irrespective of their age, ethnicity, and economic status [1]. IPV victims suffer from physical or mental health problems in the short and long term, including injury, pain, sleep problems, depression, posttraumatic stress disorder (PTSD), and suicide. Family members and children who live in or witness violence can also experience adverse health and social developmental issues [4]. Notably, children exposed to IPV are more likely to experience depression, anxiety, PTSD, dissociation, and anger [5, 6]. Physical health outcomes include injuries, sexually transmitted diseases, back and limb problems, memory loss and dizziness, and gastrointestinal conditions. Female victims could experience undesirable reproductive outcomes, including miscarriages and gynecologic disorders. From an economic perspective, the approximate IPV lifetime cost is $103,767 per female victim and $23,414 per male victim; a population economic burden amounts to nearly $3.6 trillion (2014 US$) over victims’ lifetimes, 37% of which, $1.3 trillion, is paid by the government [7].

Emerging data shows that since the outbreak of COVID-19, reports of IPV have increased worldwide. In the US, following school closures and the enactment of stay-at-home orders, the Portland Police Bureau, for example, recorded 29% and 22% increases in arrests related to domestic violence between March 12 and April 15, 2020 compared to the previous year and prior weeks, respectively [8]. Similarly, large police departments outside New York City reported a 12% increase in IPV for Q1 of 2020 compared to the same period in 2019 [9]. The current crisis, described as a “horrifying global surge in domestic violence” by the United Nations Chief [10], is attributed to mandatory lockdowns or movement restrictions to curb the spread of COVID-19 [10, 11]; isolation of IPV victims with their abusers at home; elevated tension between partners, compounded by security, health, and financial worries (e.g., socioeconomic instability and business closures) [10, 12]. Actively addressing and preventing IPV is an imperative public health agenda. Indeed, the Sustainable Development Goals—a collection of 17 interlinked global goals designed to be a “blueprint for achieving a better and more sustainable future for all—focus on eliminating all forms of violence against women and girls [13] [14].

Conventionally, IPV-related data have been collected through surveys and medical/police reports [15, 16]. It has become challenging to obtain IPV-related data from these traditional sources during the COVID-19 pandemic because IPV victims, for example, were reluctant to see healthcare providers due to the fear of the coronavirus [15, 16]. Social media websites (SMW), such as Twitter and Reddit, can potentially get around the pandemic-induced obstacles by collecting data online. More than 2.9 billion individuals use SMW globally, many of whom use it for extended periods [17]. SMWs have become a new communication tool for people to express their thoughts, emotions, opinions, and daily problems, regardless of geographical locality. Data shared over SMWs has extensive spread, diffusion, and influence. During the COVID-19 pandemic and lockdown, SMWs have become people’s primary channel to express and share information, emotions, and daily lives with their friends and family members [18]. In this regard, SMW has leverage to reach out to IPV victims because IPV victims tend to share their sensitive information more with friends (64%) and family members (49%), but less to healthcare providers (26%), police (23%), and shelter advocates (20%) [19, 20]. Also, SMW has been shown in past research to be an excellent source for collecting live data precisely, at scale (i.e., a large number of observations), anonymously (e.g., Reddit), unobtrusively, and at a low cost [21, 22]. Thus, SMW data about IPV and other digital footprints about IPV victims’ behaviors can be analyzed in real-time to model patterns of IPV and discover underlying risk factors at the population and individual levels [17]. Moreover, during the COVID-19 pandemic, the United Nations urged to increase investment in online technology and civil society organizations to build harmless means for women to find help, and support without informing their abusers and accordingly reduce domestic violence [10]. SMWs, where users can share anonymous postings, may serve as a more secure platform than other means (e.g., phone call, email) to offer instrumental, informational, and social support to IPV victims. These advantages of SMW can complement (not substitute) the conventional resources (e.g., hotline, shelter) and strengthen the effort to prevent IPV and support IPV victims.

To the best of our knowledge, there has been no study employing social media analytics (e.g., natural language processing [NLP], machine learning) to identify IPV victims on SMWs for surveillance, prevention, and/or intervention. Therefore, this pilot study aims to develop an automated, social media-based system to detect and categorize streaming IPV-related big data (into IPV and non-IPV cases) during the COVID-19 pandemic on Twitter via NLP and machine learning.

### Significance

An effective NLP pipeline, including an automatic classifier, will help develop a surveillance system for IPV self-reported on SMWs, particularly during an emergency such as the COVID-19 pandemic. Such a pipeline may be used to detect and proactively reach out to large numbers of IPV victims simultaneously to provide support, instead of waiting for them to seek help during and after the current pandemic. Also, the pipeline will lay a foundation to deliver evidence-based, non-contact interventions to IPV victims on social media for IPV prevention, mental health treatment, empowerment, and support [23].

### Contributions

- We are the first study to collect data, develop annotation guidelines, and then manually annotate a large number of tweets related to IPV.
- We are also the first to develop an effective NLP pipeline involving supervised machine learning that can automatically extract and classify IPV-related tweets.
- We present a thorough analysis of the classification errors, and the model performances at different training data sizes, as these information may be crucial for future research on improving the classifier.
- We analyzed potential biases and the trustworthiness of our model’s performance through content analysis and the approach of layered integrated gradients.
- We described challenges faced during our analysis and lessons learned, which will help future researchers design similar NLP systems for social media-based data. We also discuss the strengths and weaknesses of our model based on the analyses performed.

## METHODS

### Data collection

We collected publicly available English posts (tweets) related to IPV from Twitter using the SMW’s public streaming application programming interface (API). The keywords we used are provided in Supplementary Table S1. We also used the Python library snscrape to collect data during the COVID-19 pandemic between January 1, 2020 – and March 31, 2021, accruing a total of 1,764,707 IPV-related tweets.

### Annotation guidelines

Four annotators encoded each tweet into *personal* (reported by victims themselves) *IPV-report* (or *IPV*) or *non-IPV-report* (or *non-IPV*). Such categorization of each tweet was made based on the definition of IPV. The Centers for Disease Control and Prevention defines IPV as physical violence, sexual violence, stalking, and psychological aggression (including multiple coercive tactics) by a current or former intimate partner (i.e., spouse, boyfriend or girlfriend, dating partner, or ongoing sexual partner) [24]. Supplementary Table S2 provides comprehensive details and examples of various types of IPV. Two necessary factors that determined if a tweet was a self-reported IPV were (1) mention of intimate partner as an abuser and (2) mention or description of any type of abuse (physical violence, sexual violence, stalking, and psychological aggression) or its tactics.

The annotation was conducted iteratively over small datasets. Guided by the definition of IPV and a domain-expert (SK), the annotators discussed disagreements in the codes and reached consistent coding rules. With the finalized annotation guidelines, the annotators encoded the final dataset (n=6,348) used for training and testing our NLP model. The gold-standard dataset was developed when reliable levels of agreement between the annotators were achieved. The dataset was split into three overlapping subsets. A subset of the tweets was annotated twice or thrice for computing inter-annotator agreements (IAA). For double-annotated tweets, if the annotators disagreed with the class, the tweet was assessed by an independent annotator who resolved the disagreement. The final IAA was calculated using Cohen’s kappa [25].

### Text classification model

We investigated three different approaches for constructing an IPV-related classifier. We used four traditional machine learning algorithms: decision tree [26], support vector machine (SVM) [27, 28], random forest (RF) [29], and neural networks (NN) [30]. We also experimented with more advanced algorithms for text classification, including a deep learning-based algorithm, namely, bi-directional long short-term memory (BiLSTM) [31, 32] and two transformer-based models, namely, Bidirectional Encoder Representations from Transformers (BERT) [33, 34] and Robustly Optimized BERT (RoBERTa) [34].

#### Pre-processing stage

Before feeding the tweets to the model, they were cleaned by removing unwanted words, such as non-English characters. We replaced and anonymized the URLs and usernames with predefined tags (i.e., URL and <user>). We also lowercased all text.

#### Traditional machine learning models

For the traditional classifiers, we produced 1,000 most frequent *n*-grams (adjacent series of words with *n* ranging from 1 to 3: unigrams (*n*=1), bigrams (*n*=2), and trigrams (*n*=3)) [35].

#### Deep learning model

We converted each word into a corresponding word vector then fed it into the BiLSTM classifier. For a word to vector conversion, we used the Twitter GloVe word embeddings trained on 2B tweets and 27B tokens, including a 1.2 million-word vocabulary. We used uncased GloVe embedding with 200-dimension vectors [36].

#### Transformer-based models

We used BERT and RoBERTa. The pre-processed training tweets were fed into the model for fine-tuning the model for IPV classification. The hyper-parameters and technical details are presented in Supplementary Table S3.

#### Model training validation

Our primary objective was to create a model for identifying IPV tweets from streaming Twitter data. Our primary metric for assessing classifiers’ performance was the F1-score (harmonic mean of precision and recall) for the IPV class. We divided the annotated dataset into training (70%), development (10%), and test sets (20%). We used the training dataset for training models and the development dataset for optimizing model hyperparameters.

### Post-classification analyses

#### Learning curve analysis

We evaluated the model performance at different sizes of training data (20%, 40%, 60%, 80%, 100%) to investigate the behavior of each model at a given training percentage and assess whether growing the training dataset improved the models’ performance on the fixed test dataset.

#### Error analysis

To identify potential reasons for misclassifications, we studied the common patterns of misclassifications made by our model by manually analyzing the contents of a subset of misclassified tweets.

#### Bias analysis

Generally, humans recognize the meanings of sentences by focusing on the most important words. Some words play more important roles than others in understanding a post’s meaning and deciding its class (e.g., IPV or non-IPV). We aimed to examine the words on which our best performing model focuses and which ones have more importance than the others in making the classification decisions. This process helps us ensure that the developed model has a low bias, is trustworthy, and understands the model’s errors. To achieve this objective, we used the approach of layered integrated gradients [37] proposed by Captum [38] and [39]. The approach is based on an attribution technique called integrated gradients [40], which uses an interpretable algorithm that calculates word importance scores by approximating the integral of the gradients of the model’s outputs with respect to the inputs [40, 41].

## RESULTS

### Annotation and inter-annotator agreement

The total number of annotated tweets was 6,348, with a considerable imbalance in the distribution (non-IPV-report tweets: 5,680 [∼89%]; IPV-report tweets: 668 [∼11%]). We divided the instances via stratified sampling across training, validation, and test sets by 70%, 10%, and 20%, respectively (training 4,443, validation 635, and test 1,270). The average pairwise IAA among double-annotated tweets (N=1,834) was k=0.86 (Cohen’s kappa [25]), which can be interpreted as a substantial agreement [42].

### Classification results

The classifier performances on the test set are shown in Table 1. Accuracies and F_1_ scores are reported for each classifier. The confusion matrix for the best-performing classifier (i.e., RoBERTa) is shown in Figure 1. As Table 1 illustrates, the BERT model also obtained competitive results. However, traditional machine learning and deep learning models did not perform well, particularly for the primary development metric (IPV-report F_1_ scores), with no significant difference between these classifiers, particularly in IPV F_1_ scores.

**Table 1.** Accuracies and F_1_ scores on the test set for the traditional classifiers (decision tree, SVM, NN) and advanced transformer-based classifiers (BERT, RoBERTa).

| **Classifier** | **Accuracy (%)** | **IPV $F_1$ score** | **Non-IPV $F_1$ score** |
| --- | --- | --- | --- |
| Decision tree | 89 | 0.48 | 0.94 |
| SVM | 93 | 0.52 | 0.96 |
| Neural network | 91 | 0.50 | 0.95 |
| BiLSTM | 91 | 0.44 | 0.95 |
| Transformer (BERT) | 94 | 0.70 | 0.97 |
| Transformer (RoBERTa) | **95** | **0.76** | **0.97** |

SVM = Support Vector Machine, NN = Neural Network, BiLSTM = Bi-Directional Long Short-Term Memory, BERT = Bidirectional Encoder Representations from Transformers, RoBERTa = Robustly Optimized BERT.

**Figure 1.**
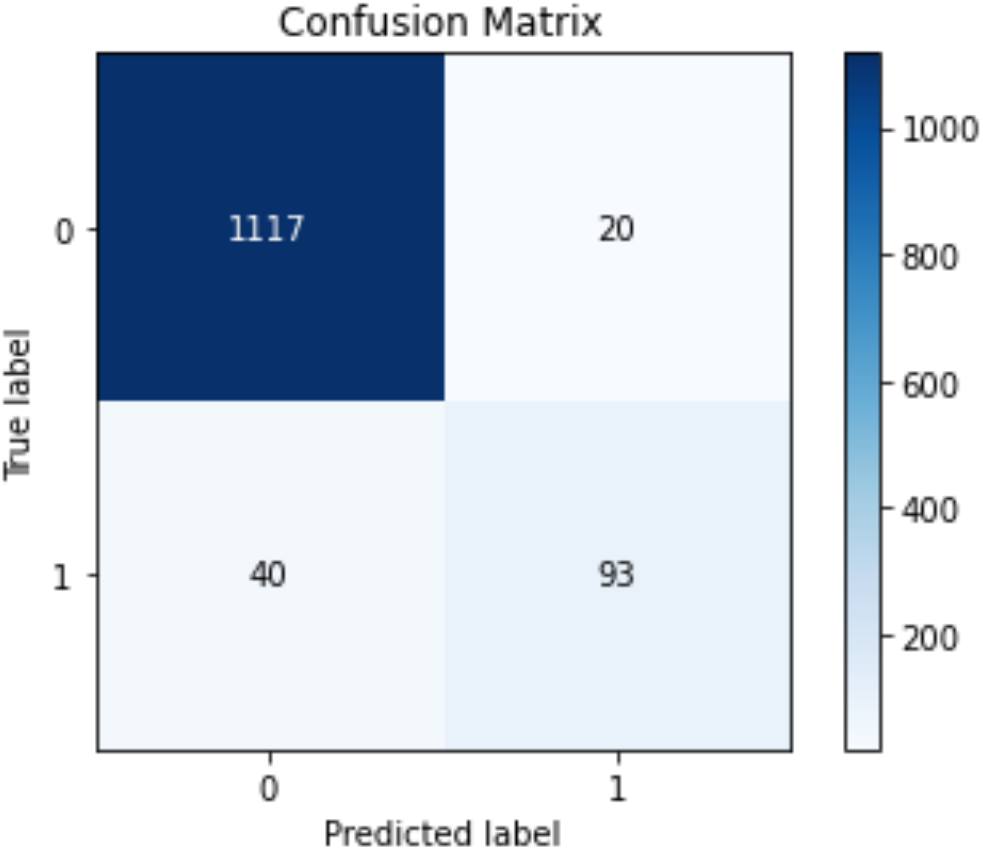
Confusion matrix for the best classifier (RoBERTa) on the IPV binary classification task.

### Learning curve

Figure 2 shows the learning curve at different percentages of training data (20%, 40%, 60%, 80%, 100%), and the performance obtailed using fixed test data. Overall, the performance tends to improve with increasing training data, which is expected, particularly from 20% to 80%. With only 40% of the training dataset, the pre-trained models (i.e., BERT, RoBERTa) performed similarly to the traditional and deep learning models with 100% of the training dataset.

**Figure 2.**
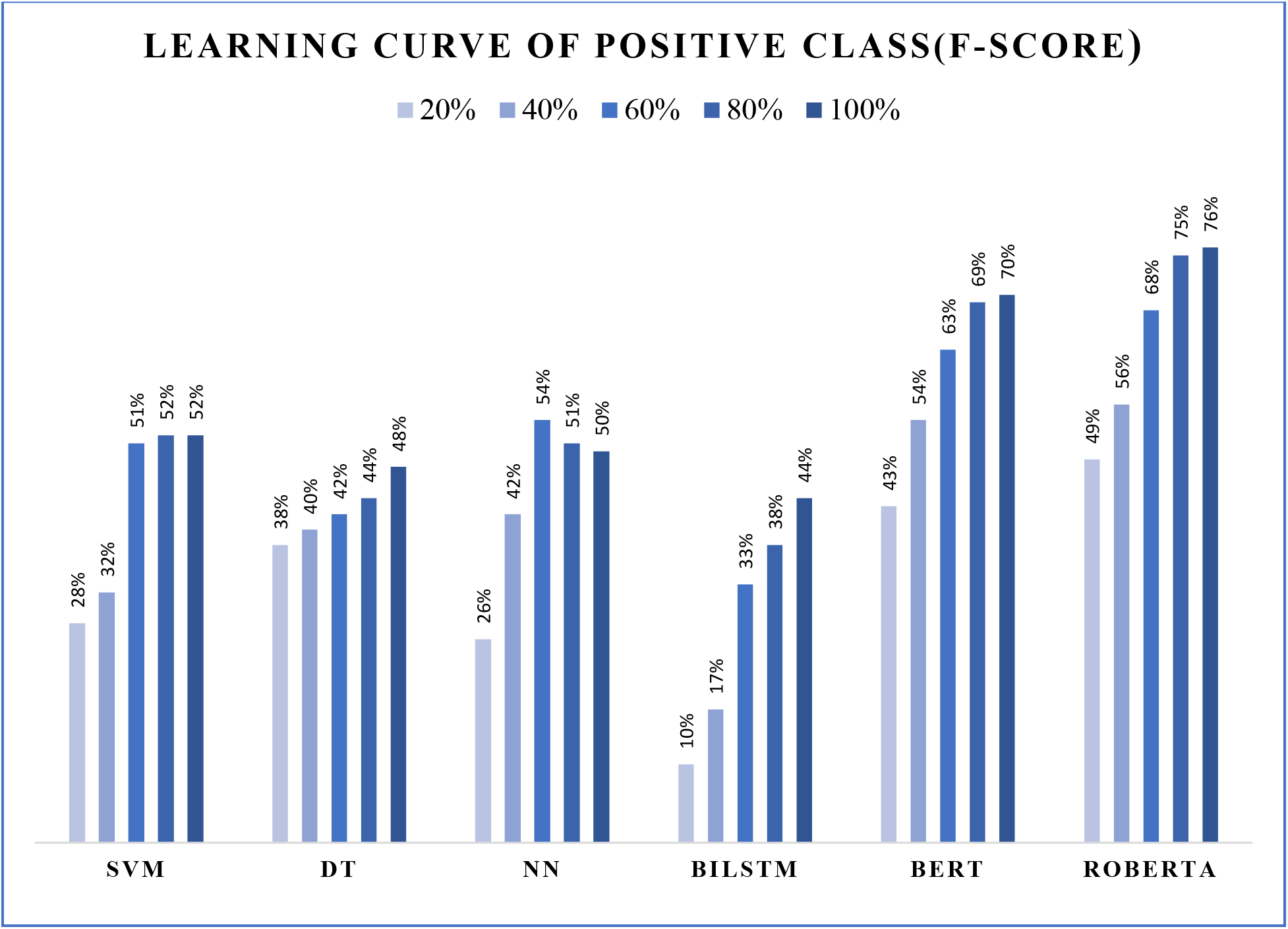
Classifier performances at different training set sizes (learning curves). SVM = Support Vector Machine, DT = Decision tree, NN = Neural Network, BiLSTM = Bi-Directional Long Short-Term Memory, BERT = Bidirectional Encoder Representations from Transformers, RoBERTa = Robustly Optimized BERT.

Given that the RoBERTa model (Table 1) produced the best performance compared with other models, we used it for our NLP pipeline, and investigated the errors made by the RoBERTa classifier. These results are presented in the following subsection.

### Error and model analyses

For the tweets annotated as *non-IPV-report*, few non-IPV examples were classified as IPV as reflected by a very high F1-score for the non-IPV class (i.e., 0.97). Nevertheless, in a few cases, an error occurred when tweets contained users’ indirect IPV experiences (reporting IPV not experienced by self but by others). For instance, “*my ex-neighbours rowed a lot. he was a big bloke, over day i heard him hit her; my heart beating out my chest, I knocked on their door. it stopped that fight. <hashtag> domestic violence*.” Additionally, non-IPV examples were misclassified as IPV when tweets contained hypothetical scenarios not experienced by the authors in the real world. One example is “*that is completely different issue. people not wanting to have children is different from divorce. i could have children with good intentions in mind, and still get divorced bc my husband turns abusive*.” In contrast, for the tweets annotated as IPV tweets, misclassification occurred into non-IPV tweets when the author’s IPV was implied in the tweet. For instance, “*I hate how abusers justify their abuse of their partner in their head. That was what my ex would do. the physical scars heal, but the mental ones remain*.” In this example, the author did not state up front that her ex-partner harmed her body and mind. “*my ex would do*” did not contain any indicator of IPV; hence, the tweet was considered to be non-IPV by our model.

### Model behavior and bias analysis

We first checked whether our model’s classification outcomes were biased toward a specific gender or racial group. We made this our focus because many recent studies have shown that machine learning models often utilize race or gender information when making inferences, rather than more relevant data [43] [44]. We selected IPV tweets that were correctly classified by our model and contained words indicating the authors’ gender (e.g., man, woman). We checked whether the model focused on male- or female-related words to make the classification decisions. Table 2 provides an example of sentences that included gender-related keywords. We used a similar approach to IPV tweets that our model misclassified. Simultaneously, we changed male-related words into female-related words and vice versa to examine if the change in the gender-based word would also alter the misclassification outcome or if this change significantly affected the word importance of the gender-based word in the tweet.

**Table 2.**
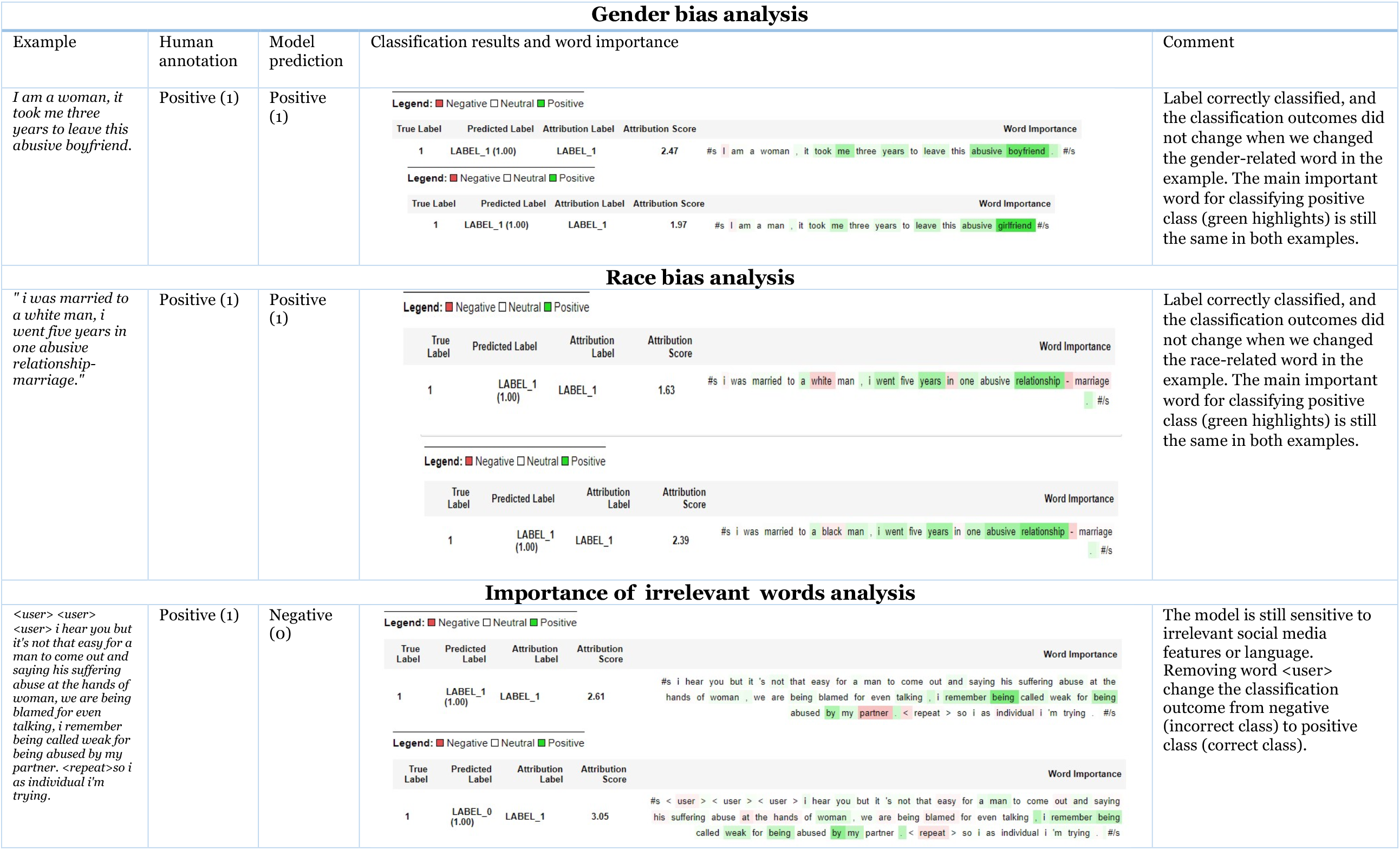
Bias and model outcome analyses were conducted using the layered integrated gradients approach. Highlighted text segments show where the model focused when making classification decisions. IPV (Positive Class), Non-IPV(Negative class)

The same method was applied to race-related words (e.g., White, Black). Table 2 shows an example. The result shows that our model’s classification decisions were not biased by gender or race. However, this analysis was executed only on a few examples, and further investigation is required in the future.

Next, we analyzed the misclassified tweets further in terms of word importance to understand the model’s focus during the classification process. We found that the model’s word importance related only slightly with the word importance focused by human annotators (an example is provided in Table 2). For instance, in the example in Table 2, the model focuses on the words “*<user>, < repeat>,”* which mean mentioning other users in the conversation and repeating the previous word, respectively. However, these features are not helpful clues for a human annotator to make the final overall classification. Importantly, however, the model did not make misclassifications based on demographic information such as race and gender.

### Use of the model on a large dataset

We were able to retrieve 153,826 tweets using IPV-specific keywords, as described in the previous section. We observed that most social media users used terms like “domestic violence” and “IPV” when discussing the topic. The hashtag “#domesticviolence” was also frequently used. Given that the main objective of the study is to identify self-reports of IPV victims, we further narrowed our pipeline by considering only those tweets which mentioned personal pronouns (“I,” “my,” “me,” “us”). Finally, 39,393 tweets from 29,583 users were collected and analyzed for content.

With the application of classification models, a total of 1,803 reports were found, comprising approximately 4.6% of the collected tweets. Further text analysis on the positive tweets showed the types of abuses that users reported and how they handled them. *N*-gram distributions helped identify the frequently discussed topics. Stop words and other commonly used terms related to this study, such as “domestic violence,” “retweet,” “behavior,” “victim,” and “violence,” were removed to reduce noise, followed by lemmatization. Obtained results show different categories of information. Primarily, most of the users described how they were victimized, and the top repeating mentions were abusive relationships (9.23%), threatening or attempting to kill (3.5%), sexual assault (2.77%), and child abuse (2.21%) (Figure 3).

**Figure 3.**
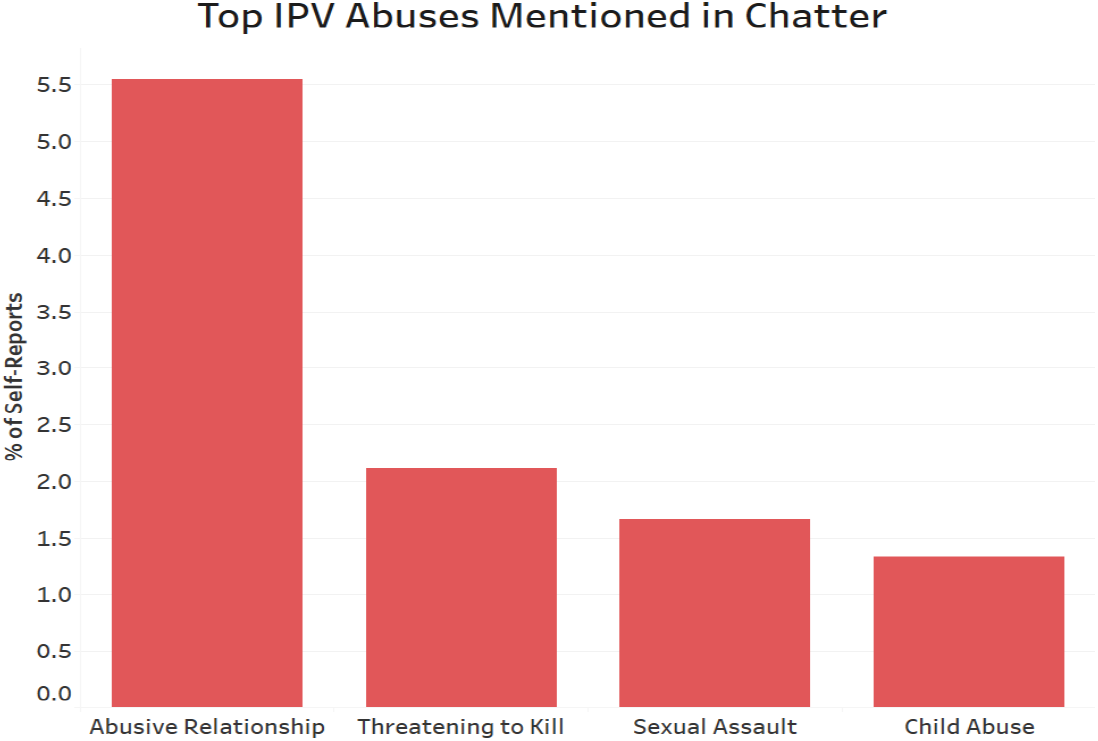
Percentage of self-reports for different types of IPV abuses (top four cases of abuse of IPV)

In contrast, as shown in Figure 4, some users expressed their personal experiences of handling violent acts, such as restraining orders (5.5%) and seeking help from police (3.3%).

**Figure 4.**
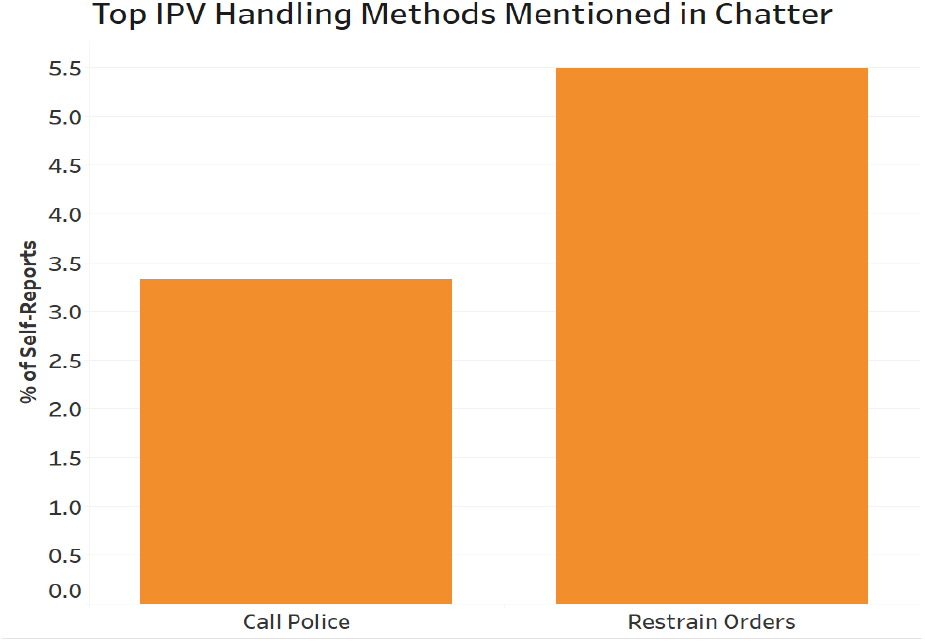
Percentage of self-reports about different ways of IPV handlings mentioned in Twitter.

A comparison of male and female self-reports shows that the number of tweets mentioning ex-husbands is three times as many as those about ex-wives. When the full dataset is analyzed (i.e., including non self-reports), mentions about ex-husbands are as much as those about ex-wives (data not shown).

## DISCUSSION

The RoBERTa classifier achieved the best performance compared with other models, and the performance was almost comparable to human annotators (based on the computed agreement of human annotators). This suggests that the developed systems can be practically employed for collecting and analyzing IPV data at a larger population level. Findings from the learning curve analysis indicate the usefulness of the per-trained model (e.g., RoBERTa) in learning with a small dataset. The pre-trained model continues improving performance with additional training data, but with a slow learning pace between 80% and 100%.

Moreover, the classification outcomes and word importance analysis show that our system is not biased to certain gender or race groups. However, this result does not necessarily mean that our developed model is bias-free, which is difficult to claim without a large test data that contains more information. Rather, we intended to ensure that the model at this stage was not making classifications based on gender- or race-related words, as well as to find the best strategies to improve the quality and diversity of training datasets in the future.

### Limitations and future directions

Based on our analysis and observations, we identified several limitations and future directions:

#### Data-centric improvement

First, the model tends to misclassify tweets that contain implicit self-report of IPV. Future work may include strategies to enrich the classification with new training datasets that address this issue. Enriching the training data can be planned to mainly boost the number of tweets with similar characteristics to those misclassified in the first training round. Second, the classifier is likely to misclassify tweets due to words and symbols that uniquely appear in social media data, such as “<,” and <user>, as well as the tweets that contain subtle language cues. Lastly, some word importance scores by our model are not highly associated with the word importance selected by a human annotator, potentially because of the data collection strategies. We used words such as “domestic violence” or “abuse” to collect our data. Therefore, positive (self-report of IPV) and negative (non-self-report of IPV) examples in our training and test data have similar distributions of these words. Thus, the model does not give these words high importance. Furthermore, although such strategies are essential only to extract relevant tweets, we are likely to miss the IPV tweets that do not contain any of our IPV-related words.

Even though our model does not show any observable bias toward gender and race while making its classification outcomes, further examination is required before and during the deployment stage to keep the model unbiased and trustworthy and prevent any data drift that may cause undetectable biases. Although not identified through content analysis, we suspect insufficient context of the events as a possible reason for misclassifications. Twitter as a micro-blogging platform allows only a limited number of characters in one tweet. Therefore, users often attempt to embed all their experiences in short sentences, forgoing detailed context that would be useful for accurate classification into either IPV or non-IPV. Along the same line, one tweet lacks context because it could be part of a series of tweets (thread) explaining the IPV experience. Automatic classifiers do not have as much an ability as humans to infer the whole meaning from a fraction of the data or by piecing information together.

### Model improvement

The dataset used to pre-train the RoBERTa or the BERT model mainly contains a corpus of books (800 million words) and English Wikipedia. The way people express themselves in social media language is different from that in books [35]. Efforts have been made to pre-train BERT-related models on social media. However, previous research showed that the RoBERTa model based on traditional text still performs better than these models in several social media NLP tasks [45] [46]. A potential direction may be combining traditional data with social media data to have a model built with a large text corpus and social media language.

## CONCLUSION

Although IPV victims often reach out for support and intervention through social media channels such as Twitter, there is little effort to use such platforms to address this public health problem. Posts about IPV are typically lost in the massive volume of data constantly posted on social media. Developing an effective, low-biased, and trustworthy model for classifying self-reported IPV in social media has significant practical applications. This study developed and evaluated an NLP pipeline to collect and classify posts from the Twitter platform to identify IPV-related tweets. Our NLP pipeline achieved comparable performance to humans, and was shown to be particularly not biased to gender or race-related words. By identifying IPV victims on Twitter, our model will lay the groundwork to design and deliver evidence-based interventions and support to IPV victims, and potentially enable us to address the problem of IPV in close to real-time via social media.

## Data Availability

Data will not be made publicly available due to the sensitive nature of the study. Interested researchers will be provided data following the execution of a data use agreement.

## Notes

### Competing Interest Statement

The authors have declared no competing interest.

### Funding Statement

The study was funded by the Injury Prevention Research Center at Emory (IPRCE), Emory University.

### Author Declarations

Emory University Institutional Review Board

