## Supplementary material for "Natural Language Model for Automatic Identification of Intimate Partner Violence Reports from Twitter"

1- **Data collection keywords**

**Table S1**. The list of keywords for data collection using Twitter API and the list of text pattern for filtering

| Keywords for data collection | Text pattern for filtering |
| --- | --- |
| *relationship*  *abusive*  *domestic*  *husband*  *partner*  *violence*  *violent*  *intimate*  *[#]ipv*  *[#]domesticabuse*  *[#]dv* | *abusive.*relation*  *domestic.*abuse*  *abusive.*husband*  *domestic.*violence*  *intimate.*partner.*violence*  *partner.*violent*  *partner.*violence*  *#ipv*  *#domesticviolence*  *#intimatepartnerviolence*  *#dv* |

2- **Annotation guidelines**

1. IPV-related vs. Not

It will be determined based on the definition of various types of intimate partner violence (IPV).

IPV includes physical violence, sexual violence, stalking, and psychological aggression (including coercive tactics) by a current or former intimate partner (i.e., spouse, boyfriend/girlfriend, dating partner, or ongoing sexual partner).

Must read: <https://www.cdc.gov/violenceprevention/intimatepartnerviolence/resources.html> (p. 11-15)

**Two necessary factors to be considered as IPV are (1) mention of intimate partner as an abuser and (2) mention/description of any types of abuse (physical violence, sexual violence, stalking, and psychological aggression)

| Category 1 | Category 2 | Example |
| --- | --- | --- |
| IPV-related | Intimate partner | - ‘My abusive ex,’ ‘My husband,’ ‘My boyfriend’ … - ‘injured partner’ |
|  | General IPV indicator | - ‘I * abusive relationship’ - ‘(intimate partner) do me serious harm’ - ‘I begged,’ ‘I pleaded’ - ‘cry’ - ‘I've been through counseling to deal with what happened.’ |
|  | Domestic violence combined with specific indicator(s) | - General violence + 'another victim' + purple heart 💜  (IPV symbol) - Domestic violence victim + purple emoji (e.g., ☯️ ) - Domestic violence victim + ‘my baby’ - Domestic violence + supportive family and friends + no mention of partner (e.g., I should’ve listened to my family and friends bro, my souvenir was a domestic violence police report and a dozen bruises) - Domestic violence + stalking – stalking from a family member is less likely - Domestic violence + male victim abused by female |
|  | Physical violence | ‘physical abuse,’ ‘physically,’ ‘physically abused’  ‘attack’  ‘beat,’ ‘beaten,’ ‘beating’  ‘bruises,’ ‘black eyes’  ‘choke,’  ‘concussion’  ‘crack’  ‘grabbing’  ‘head injuries from my (intimate partner)’  ‘hit’  ‘hold (body part(s))’ (e.g., hold the back of my neck)  ‘kicked’  ‘tried to kill me’  ‘limp’ (victim or pet)  ‘pain’  ‘post concussive syndrome’  ‘punch (body part (s))’  ‘push (body part (s))’  ‘put the shower head in my mouth’  ‘restraining’  ‘shove’  ‘smash’  ‘strangle’  ‘suffocate’ |
|  | Sexual violence | ‘sexual abuse,’ ‘sexual coercion’  ‘convincing after saying no’ |
|  | Stalking | ‘stalk’  ‘showed up outside my house again’ |
|  | Psychological aggression | ‘mental abuse’  ‘emotional abuse,’ ‘emotionally abused’  ‘verbal abuse,’ ‘verbally abused’  ‘hurtful,’ ‘mean,’ ‘stomping all over your feelings’    1) Expressive aggression  ‘call me *’ (e.g., pig, evil person, selfish, heartless bitch),  ‘name calling’  ‘insults’  ‘scream’    ‘blaming me,’ ‘saying that I was responsible for whatever happened to him,’ ‘he said that this was my fault,’ ‘making you feel like you're doing something wrong,’ ‘put me down and make me feel less, I wasn't good enough’      2) Coercive control  ‘made me watch punching my pet’  ‘forcing me’  ‘not allowed *,’ ‘wouldn’t let me *’  ‘denying access to medical care’  ‘* restriction’ (e.g., sleep, food, medicine)  ‘controlled,’ ‘controlling,’ ‘random phone and computer history checks,’ ‘scrutinized,’ ‘reads all texts,’ ‘tracks her location,’ ‘financial control,’ ‘kept a camera on me 24/7’  ‘cut off all my friends,’ ‘isolated’  ‘he grabbed a plastic bag and put it over his face,’ ‘he made me put him in a bag and put him in the ground’    3) Threat of physical or sexual violence  ‘threats,’ ‘threatened to kill *,’ ‘he said that when we get home he used to be the s*** out of me’  ‘held weapons’    4) Exploitation of victim’s vulnerability  ‘denying access to medical care’  ‘Whenever we have a disagreement, he loves to say that being married was his compromise and that he “protected” my reputation so the least I could do is give him this one thing’  ‘he tries to use it to get his way’    5) Gaslighting  ‘gaslighting’  ‘manipulation’ |
| Not related  (Not including description of self or other’s personal IPV experience) | Abuse not by an intimate partner | ‘my dad’  ‘my sister’  ‘my brother hit my mother’  ‘roommate’  ‘abusers’ - plural + no mention of intimate partner |
|  |  | ‘Abusive home,’ ‘abusive household,’ ‘abused as a child,’ ’domestic violence I grew up in’ |
|  | Hypothetical | If he was, imagine |
|  | Article (magazine, news, journal) | |
|  | Quote from books, celebrities, social media, etc. | ‘adapted,’ ‘excerpted, ‘Instagram,’ ‘notes from,’ ‘from’ |
|  | Depiction/discussion about abusive relationships in the media (e.g., movie) | |
|  | In the context of (video) games (e.g., Animal Crossing) | |

2. Self-report vs. Report from family/friend

| Self-report | [From original posts]  ‘I,’ ‘me,’ ‘my,’ ‘we,’ ‘us’  ‘my abusive relationship’  ‘I’m in an abusive relationship’  ‘when I left my abuser,’ ‘I left my husband’  ‘I ran for our lives’    [From response thread]  ‘I know *’ (e.g., how much it hurts)  ‘I relate to’  ‘* is my life’  ‘I experienced’  ‘same situation,’ ‘same happened to me,’ ‘I can soon do the same,’ ‘I did the same thing,’ ‘same here,’ ‘like me’  ‘I went through the exact same kinds’  ‘Your situation sounds like mine’  ‘This is exactly what my (intimate partner) was like’  ‘trust me’ |
| --- | --- |
| Report from family/friend | ‘my friend’  ‘my husband’s sister’  ‘her boyfriend’ ‘her husband’ ‘his wife’  ‘she’  ‘sister’  ‘my neighbor’ |

3- **Model hyperparameters**

Table S3. The hyperparameters for the classification experiments

| *Algorithm* | *Hyperparameters* |
| --- | --- |
| ***DT*** | *criterion='gini', splitter='best', max_depth=None, min_samples_split=2* |
| ***NN*** | *solver='lbfgs'*  *alpha=1e-5*  *hidden_layer_sizes=(5,2)* |
| ***SVM*** | *C parameter: 16*  *kernel: rbf*  *penalty: L1 regularization* |
| ***BLSTM*** | *Tokenization: num_words: 20727 unique tokens , maxlen: 128*  *Word Embeddings:Twitter Glove, 200 dimensional vectors*  *optimizer: ‘adam’*  *dropout rate: 0.2*  *recurrent dropout: 0.2*  *metrics: accuracy*  *epochs: 100* |
| BERT | max_seq_lenth: 128  Model: Bert_large  train_batch_size: 8  num_train_epochs: 6  learning_rate: 1e-5 |
| RoBERTa | max_seq_lenth: 128  Model: RoBERTa _large  train_batch_size: 8  num_train_epochs: 6  learning_rate: 1e-5 |
